# Prevalence, Comorbidity, and Sociodemographic Correlates of Psychiatric Disorders in the All Of Us Biobank

**DOI:** 10.1101/2021.12.02.21266700

**Authors:** Peter B. Barr, Tim B. Bigdeli, Jacquelyn M. Meyers

**Affiliations:** Department of Psychiatry and Behavioral Sciences, SUNY Downstate Health Sciences University, Brooklyn, NY; VA New York Harbor Healthcare System, Brooklyn, NY

## Abstract

**Importance:** All of Us is a landmark initiative for population-scale research into the etiology of psychiatric disorders and disparities across various sociodemographic categories.

**Objective:** To estimate the prevalence, comorbidity, and demographic covariates of psychiatric and substance use disorders in the All of Us biobank.

**Design, Setting, and Participants:** We estimated prevalence, overlap, and demographic correlates for psychiatric disorders derived from electronic health records in the All of Us biobank (release 5; N = 331,380)

**Exposures:** Social and demographic covariates.

**Main Outcome and Measures:** Psychiatric disorders derived from ICD10CM codes and grouped into phecodes across six broad domains: mood disorders, anxiety disorders, substance use disorders, stress-related disorders, schizophrenia, and personality disorders.

**Results:** The prevalence of various disorders ranges from approximately 15% to less than 1%, with mood and anxiety disorders being the most common, followed by substance use disorders, stress-related disorders, schizophrenia, and personality disorders. There is substantial overlap among disorders, with a large portion of those with a disorder (~57%) having two or more registered diagnoses and tetrachoric correlations ranging from 0.43 – 0.74. The prevalence of disorders across demographic categories demonstrates that non-Hispanic whites, those of low socioeconomic status, women and those assigned female at birth, and sexual minorities are at greatest risk for most disorders.

**Conclusions and Relevance:** Although the rates of disorders in All of Us are lower than rates for disorders in the general population, there is considerable variation, comorbidity, and differences across social groups. Large-scale resources like All of Us will prove to be invaluable for understanding the causes and consequences of psychiatric conditions. As we move towards an era of precision medicine, we must work to ensure it is delivered in an equitable manner.

## INTRODUCTION

Psychiatric and substance use disorders remain an ever-present challenge to public health, incurring significant cost to society, affected individuals, and their families. Mental illness is estimated to contribute ~13% to the global burden of disease^1^ and the human cost can be staggering: with over 49,000 Americans dead as the result of an opioid overdose and over 47,000 deaths attributable to suicide in 2019, alone^2^. Psychiatric disorders incur significant financial cost, with estimates in the United states ranging from ~$320 billion annually for serious mental illnesses^3^ to ~$78 billion annually for opioid use^4^. Importantly, these disorders rarely manifest in isolation, showing strong patterns of comorbidity and shared risk factors^5–7^.

Historically, estimating the prevalence of psychiatric disorders has relied on nationally representative or population-based surveys often conducted by trained lay interviewers^8–10^, or through clinical ascertainment^11^. More recently, large-scale biobanks and healthcare systems are becoming increasingly used for research purposes. To date, resources such as the UK Biobank^12^, the Million Veteran Program^13^, Biobank Japan^14^, and FinnGen, among others, have proved to be an invaluable resource in the understanding of the causes of various psychiatric and related disorders. While valuable, these resources may not reflect the unique context within the US. Additionally, much of the research conducted in this area has focused primarily on individuals of European ancestries^15,16^. This is despite the previously observed differences in rates and patterns of psychiatric disorders by race-ethnicity, as well as known risk factors that are especially pertinent in the US given its sordid history with racism and inequality. In order to improve diversity in health-focused epidemiological research, further interrogate existing health disparities, and deliver on the promise of equitable precision medicine, the National Institutes of Health launched the All of Us program, a historic effort to collect and study data from at least one million people living in the United States^17^. Beginning in 2018, participants were able to complete surveys, provide genotypic data, and link their electronic health records to help build a comprehensive database of their health-related data. The goal of the All of Us program is to use this data to understand how biology, lifestyle and social determinants come together to affect health, and ultimately to treat and prevent illness.

The current analysis focuses on characterizing the prevalence, comorbidity, and sociodemographic disparities for psychiatric and substance use disorders in the All of Us database. We compare these estimates to those from nationally representative and population-based samples to help better understand the etiology of psychiatric disorders in All of Us, and the participants that make up this new sample.

## METHODS

### The All of Us Biobank

The All of Us Research Program is a prospective cohort study aiming to recruit at least one million individuals in the United States, with the overall goal of providing a unique resource to study the effects of lifestyle, environment and genomics on health and health outcomes. Participant recruitment is predominantly done through participating health care provider organizations and in partnership with Federally Qualified Health Centers, with an emphasis on recruiting persons affiliated with those centers. Interested potential participants can also enroll in the program as direct volunteers, visiting community-based enrollment sites. Initial enrollment, informed consent (including consent to share EHRs), and baseline health surveys are done digitally through the All of Us program website (https://joinallofus.org). Once this step is completed, the participant is invited to undergo a basic physical exam and biospecimen collection at the affiliated healthcare site. Participant follow-up is done in two ways, passively via linkage with EHR and actively by periodic follow-up surveys. For this study, we included data from participants enrolled in the study between May 6, 2018 and April 1st, 2021 (All of Us release 5, N = 331,380). This work was performed on data collected by the previously described All of Us Research Program using the All of Us Researcher Workbench, a cloud-based platform where approved researchers can access and analyze All of Us data. All analyses were in concordance with the ethical guidelines outlined in the All of Us Code of Conduct.

### Psychiatric Diagnoses

We derived diagnoses based on phecodes from registered ICD10CM codes in the All of Us system. In the case where participants had multiple documentations of the same code, we used the most recent instance. First, we selected all relevant ICD codes for the more prevalent psychiatric disorders, including mood, anxiety, substance use (excluding nicotine), stress-related, psychotic, personality, eating, and other disorders. We then clustered these ICD codes using phecodes^18^ from the *PheWAS* package in R^19^. Phecodes provide a convenient way to cluster codes in EHR based data^20^. A full list of the EHR descriptions, ICD10CM codes, and corresponding phecodes is provided in eTables 1 and 2 in the supplement.

### Sociodemographic Risk Factors

All of Us provides extensive measures of social and demographic characteristics. Our analyses included covariates for age, sex assigned at birth (male, female, and no answer), gender identity (man, woman, or no answer), sexual orientation (straight/heterosexual vs. all others), race-ethnicity (non-Hispanic white, Black/African-American, Hispanic or Latino/a/x, Asian, other race-ethnicity, and multiracial), educational attainment (less than HS, HS or equivalent, some college, and college degree or more), household income (ranges of categories from under $25K to $100K or more annually), access to health insurance, and country of origin (US vs foreign born) Given previous research on the widespread mental health disparities across these social factors^8,21^, it is important to assess whether there are similar patterns in the All of Us data to ensure that it accurately reflects the broader US population.

### Analytic Strategy

Our analytic strategy was threefold. First, after cleaning and collating the EHR data, we estimated the baseline lifetime prevalence of each disorder, comparing this prevalence to estimate of population lifetime prevalence (where available) from other published reports of nationally representative or similar data. Second, we estimated the conditional odds ratios (OR) using logistic regression models with each of the social and demographic covariates included. To correct for the number of independent models, we used a Bonferroni correction (*p* < .05/6 = .0083) Finally, we calculated the cooccurrence of each of the disorders and estimated the tetrachoric correlations to characterize overlap in disorders and patterns of comorbidities.

## RESULTS

### Sample Characteristics and Prevalence of Psychiatric Disorders

Table 1 presents the sample demographic characteristics. All of Us included questions for sex assigned at birth (38.1% Male, 60.7% Female, 1.3% Neither/skipped) and gender identity (37.8% Man, 60.2% Woman, 2.1% Neither/skipped). The vast majority of respondents identified as heterosexual (~89%). The sample is highly educated, with approximately 40% of the sample reporting a college degree or higher. However, when we compare this distribution to household income, we see that there is considerably more variation, with ~20% reporting an annual household income below $25K and ~20% reporting an annual household of $100K or more (though ~20% skipped this question). One of the goals of All of Us was to recruit a more racially and ethnically diverse sample, and this is reflected in the sample compositions (52% non-Hispanic White, 21% Black/African American, 18% Hispanic or Latino/a/x, 3% Asian, and 3% who identify as some other race-ethnicity or multiracial). Most participants were born in the United States (~84%).

**Table 1:** All of Us Sample Characteristics (N = 329,038)

|  |  | **N** | **%** |
| --- | --- | --- | --- |
| Sex assigned at birth | Female | 199,658 | 60.68% |
|  | Male | 125,184 | 38.05% |
|  | Not male, not female, no matching concept, prefer not to answer, or skipped | 4,196 | 1.28% |
| Gender | Woman | 197,961 | 60.16% |
|  | Man | 124,202 | 37.75% |
|  | Not man only, not woman only, no matching concept, prefer not to answer, or skipped | 6,875 | 2.09% |
| Sexual orientation | Straight/Heterosexual | 290,176 | 88.19% |
|  | All others | 38,862 | 11.81% |
| Age | 18-29 | 48,438 | 14.72% |
|  | 30-44 | 74,353 | 22.60% |
|  | 45-64 | 128,463 | 39.04% |
|  | 65+ | 77,784 | 23.64% |
| Race-ethnicity | Non-Hispanic White | 172,753 | 52.50% |
|  | Black/African American | 69,084 | 21.00% |
|  | Hispanic or Latino/a/x | 59,348 | 18.04% |
|  | Asian | 10,747 | 3.27% |
|  | Other race-ethnicity | 5,697 | 1.73% |
|  | Multiracial | 5,282 | 1.61% |
|  | Missing | 6,127 | 1.86% |
| Education | Less than high school | 32,178 | 9.78% |
|  | High school or equivalent | 65,214 | 19.82% |
|  | Some college | 83,518 | 25.38% |
|  | College grad/advanced degree | 140,542 | 42.71% |
|  | Missing | 7,586 | 2.31% |
| Household Income | Less than 25k | 90,190 | 27.41% |
|  | 25k to 50k | 49,190 | 14.95% |
|  | 50k to 75k | 33,696 | 10.24% |
|  | 75k 100k | 25,867 | 7.86% |
|  | more than 100k | 66,389 | 20.18% |
|  | Missing | 63,706 | 19.36% |
| Health insurance | Yes | 296,627 | 90.15% |
|  | No | 23,235 | 7.06% |
|  | Missing | 9,176 | 2.79% |
| Marital status | Married | 137,597 | 41.82% |
|  | Living With Partner | 21,906 | 6.66% |
|  | Divorced | 46,496 | 14.13% |
|  | Never Married | 85,479 | 25.98% |
|  | Separated | 11,655 | 3.54% |
|  | Widowed | 16,779 | 5.10% |
|  | Missing | 9,126 | 2.77% |
| Birthplace | USA | 275,464 | 83.72% |
|  | Foreign born | 50,237 | 15.27% |
|  | Missing | 3,337 | 1.01% |

Table 2 presents lifetime prevalence for psychiatric disorders. Within the All of Us EHR data, mood disorders are the most common (14.31%), with major depressive disorder being the most common form of mood disorder (12.61%), followed by bipolar disorder (3.21%), and dysthymic disorder (0.89%). Followed closely in prevalence to mood disorders, are those with any anxiety disorder (13.87%). Within anxiety disorders, anxiety disorder (typically unspecified) is the most prevalent (12.86%), followed by generalized anxiety disorder (3.87%), social anxiety disorder/agoraphobia (1.63%), and other phobias (0.38%). In terms of substance use disorder (SUD), 5.91% have a documented SUD, with 2.96% having a documented alcohol use disorder (AUD) and 4.35% having a documented SUD related to some other form of substance (see eTable 3 for breakdown by substance). For stress related disorders, 2.6% had a documented adjustment disorder and 2.32% received a post-traumatic stress disorder (PTSD) diagnosis, and a total of 4.54% have any documented stress-related disorder. Roughly 2% of All of Us participants have a documented sleep disorder. Attention-deficit hyperactivity disorder (ADHD) and schizophrenia had an estimated prevalence of ~1%, Personality disorders, eating disorders, and obsessive-compulsive disorder each had a prevalence below 1%. In almost every case, the prevalence of these disorders is lower than the prevalence estimated using nationally representative samples^6,8,21–27^.

**Table 2:**
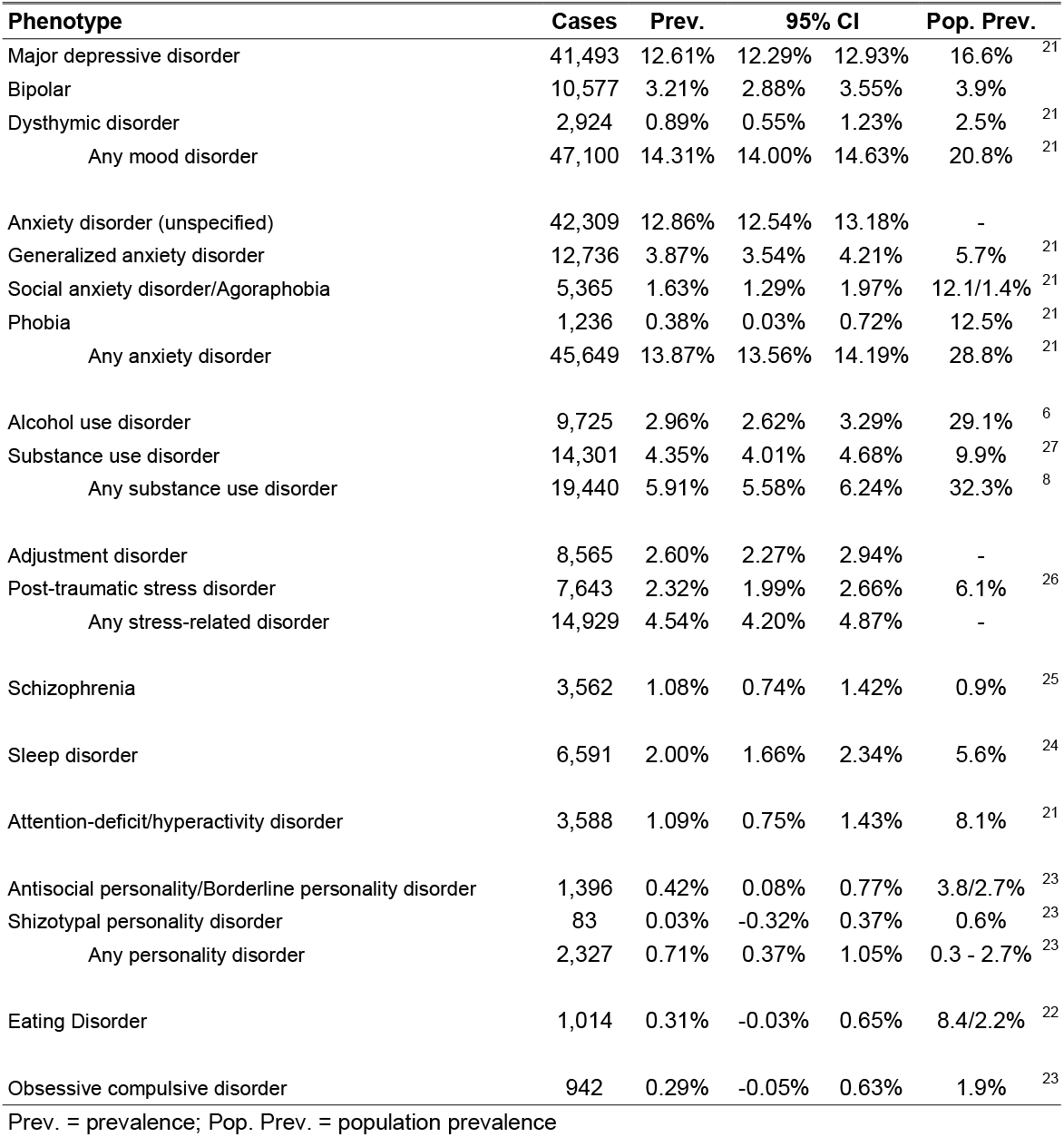
Prevalence of Psychiatric Diagnoses in the All of US EHR (N = 329,038)

| Phenotype | Cases | Prev. | 95% CI |  | Pop. Prev. |  |
| --- | --- | --- | --- | --- | --- | --- |
| Major depressive disorder | 41,493 | 12.61% | 12.29% | 12.93% | 16.6% | <sup>21</sup> |
| Bipolar | 10,577 | 3.21% | 2.88% | 3.55% | 3.9% |  |
| Dysthymic disorder | 2,924 | 0.89% | 0.55% | 1.23% | 2.5% | <sup>21</sup> |
| Any mood disorder | 47,100 | 14.31% | 14.00% | 14.63% | 20.8% | <sup>21</sup> |
| Anxiety disorder (unspecified) | 42,309 | 12.86% | 12.54% | 13.18% | - |  |
| Generalized anxiety disorder | 12,736 | 3.87% | 3.54% | 4.21% | 5.7% | <sup>21</sup> |
| Social anxiety disorder/Agoraphobia | 5,365 | 1.63% | 1.29% | 1.97% | 12.1/1.4% | <sup>21</sup> |
| Phobia | 1,236 | 0.38% | 0.03% | 0.72% | 12.5% | <sup>21</sup> |
| Any anxiety disorder | 45,649 | 13.87% | 13.56% | 14.19% | 28.8% | <sup>21</sup> |
| Alcohol use disorder | 9,725 | 2.96% | 2.62% | 3.29% | 29.1% | <sup>6</sup> |
| Substance use disorder | 14,301 | 4.35% | 4.01% | 4.68% | 9.9% | <sup>27</sup> |
| Any substance use disorder | 19,440 | 5.91% | 5.58% | 6.24% | 32.3% | <sup>8</sup> |
| Adjustment disorder | 8,565 | 2.60% | 2.27% | 2.94% | - |  |
| Post-traumatic stress disorder | 7,643 | 2.32% | 1.99% | 2.66% | 6.1% | <sup>26</sup> |
| Any stress-related disorder | 14,929 | 4.54% | 4.20% | 4.87% | - |  |
| Schizophrenia | 3,562 | 1.08% | 0.74% | 1.42% | 0.9% | <sup>25</sup> |
| Sleep disorder | 6,591 | 2.00% | 1.66% | 2.34% | 5.6% | <sup>24</sup> |
| Attention-deficit/hyperactivity disorder | 3,588 | 1.09% | 0.75% | 1.43% | 8.1% | <sup>21</sup> |
| Antisocial personality/Borderline personality disorder | 1,396 | 0.42% | 0.08% | 0.77% | 3.8/2.7% | <sup>23</sup> |
| Shizotypal personality disorder | 83 | 0.03% | -0.32% | 0.37% | 0.6% | <sup>23</sup> |
| Any personality disorder | 2,327 | 0.71% | 0.37% | 1.05% | 0.3 - 2.7% | <sup>23</sup> |
| Eating Disorder | 1,014 | 0.31% | -0.03% | 0.65% | 8.4/2.2% | <sup>22</sup> |
| Obsessive compulsive disorder | 942 | 0.29% | -0.05% | 0.63% | 1.9% | <sup>23</sup> |

Prev. = prevalence; Pop. Prev. = population prevalence

### Association of Social and Demographic Covariates with Psychiatric Disorders

Table 3 includes the estimates for the associations between demographic covariates and six categories of psychiatric disorders: any mood disorder (MOOD), any anxiety disorders (ANX), any substance use disorder (SUD), any stress-related disorders (STRESS), schizophrenia (SCZ), and any personality disorder (PERS).

**Table 3:**
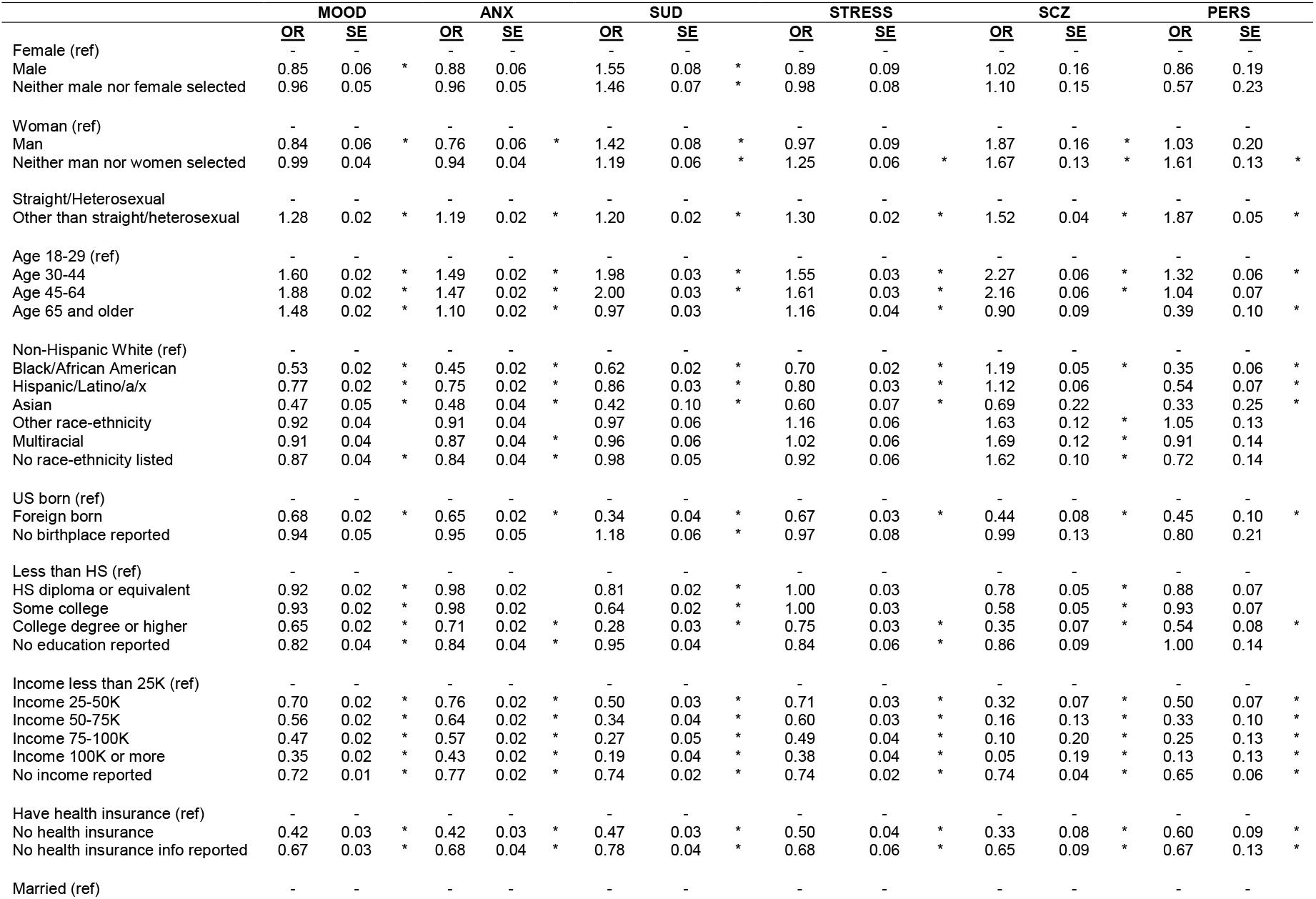
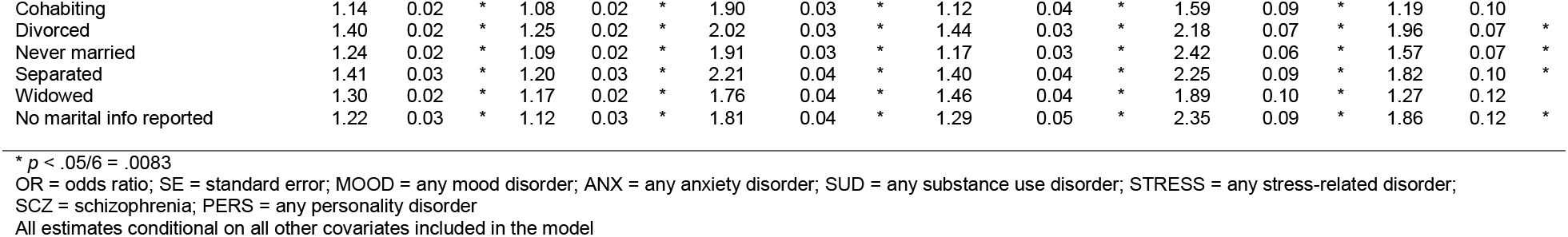
Adjusted Estimates of Risk for Psychiatric Disorders.

Similar patterns emerge across sex and gender, where those assigned male at birth and those who identify as men have lower odds of mood (OR_SEX_ = 0.85; OR_GENDER_ = 0.84) and anxiety (OR_SEX_ = 0.88; OR_GENDER_ = 0.75) disorders, but increased risk for SUDs (OR_SEX_ = 1.55; OR_GENDER_ = 1.42) and schizophrenia (OR_GENDER_ = 1.87). Those who did not identify with hegemonic categories for sex assigned at birth or gender, and those who did not identify as straight/heterosexual were at increased risk across SUDs (OR_SEX_ = 1.46; OR_GENDER_ = 1.19; OR_SEX. ORIEN._ = 1.20), stress-related disorders (OR_GENDER_ = 1.25; OR_SEX. ORIEN._ = 1.30); schizophrenia (OR_GENDER_ = 1.67; OR_SEX. ORIEN._ = 1.52), and personality disorders (OR_GENDER_ = 1.61; OR_SEX. ORIEN._ = 1.87).

Compared to those aged 18 to 29, older individuals are at increased risk for almost all of the disorders included in this analysis, with odds ratios ranging from 1.10 to 2.27. The exception is for those aged 65 and older being at reduced risk for personality disorders (OR = 0.39). Compared to Non-Hispanic White participants, individuals of all other racial ethnic categories are at reduced risk for mood, anxiety, substance use, stress-related, and personality disorders, to varying degree. However, for schizophrenia, Black/African American (OR = 1.19) participants, those of other racial-ethnic groups (OR = 1.63), and those who identify as multiracial were all at increased risk (OR = 1.69). Those who were born outside the US were at reduced risk for every type of disorder (ORs = 0.34 – 0.68).

Socioeconomic variation provides a more complicated picture. Those with a college degree or higher have lower risk of all six categories of disorder relative to those with less than a high school education (ORs = 0.35 – 0.71), but those with some college or the equivalent of a high school diploma only have reduced risk of mood disorders, SUDs, and schizophrenia, and the effect sizes tended to be smaller (ORs = 0.58 – 0.93). For income, however, individuals in *any* income category have a relatively lower risk of psychiatric disorder across any of the six categories (ORs = 0.05 – 0.70). Those reporting no health insurance had much lower odds of having any disorder (ORs = 0.33 – 0.60), which likely reflect their inability to access care. Those who are never married, cohabiting, separated, divorced, or widowed had much higher risk of every class of disorder (ORs = 1.08 – 2.21).

Figure 1 presents the prevalence by each of the demographic categories included in the analyses, above (see supplemental S4 for exact values). The differences across groups align almost entirely with the results from the conditional models, with a few notable exceptions. The raw prevalence for any SUD is ~8% in among those who are Black/African American, compared to ~ 5% in those who are non-Hispanic White, the opposite pattern seen in the conditional analyses. The reversed direction of association for Black/African Americans, from being at increased risk in bivariate associations to being at reduced risk in the conditional models, is accounted for by the inclusion of educational attainment, household income, and health insurance, suggesting that much of the risk is related to socioeconomic inequalities.

**Figure 1:**
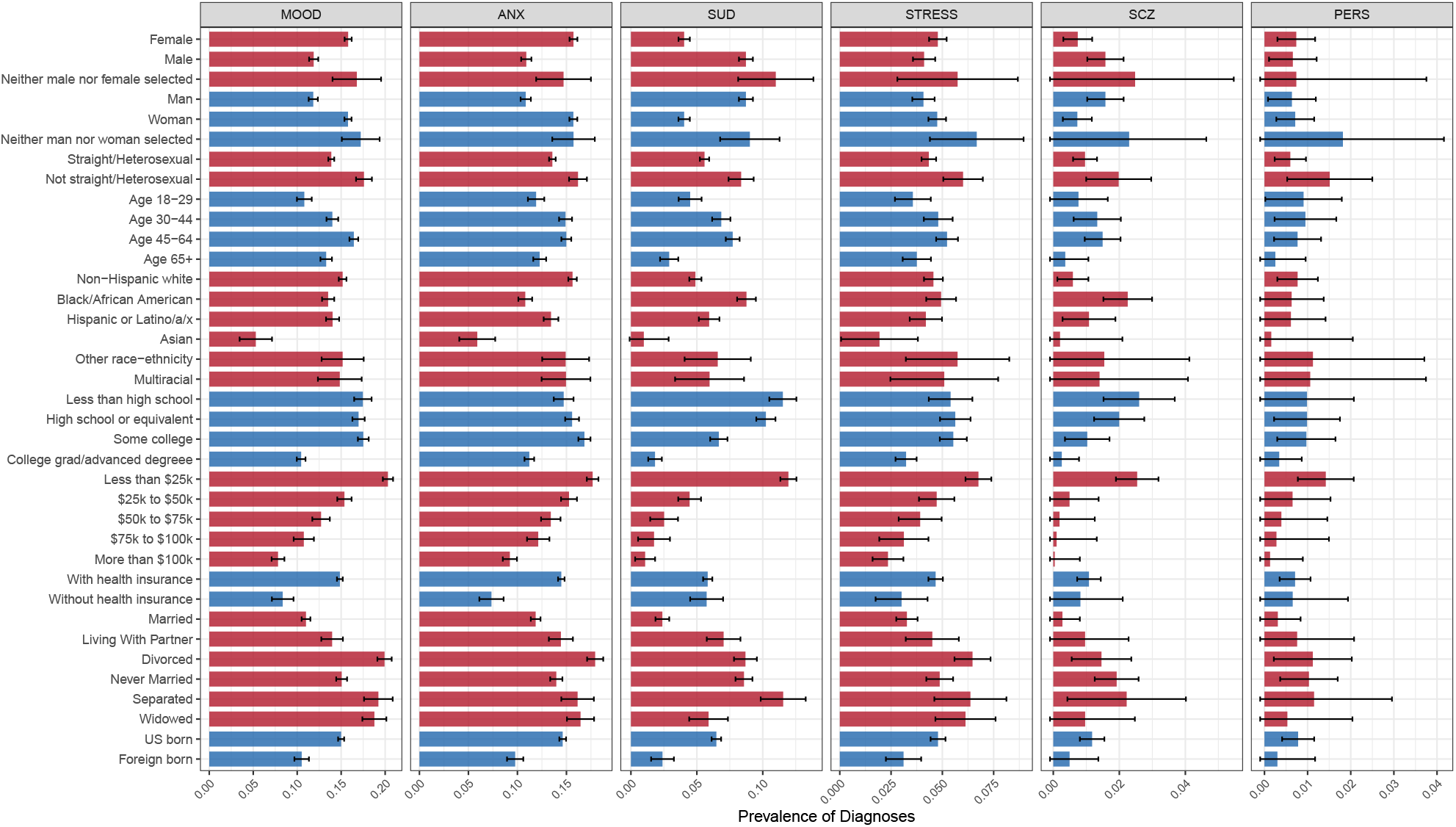
Prevalence of Psychiatric Disorders across Social and Demographic Covariates. Unconditional prevalence of each category of psychiatric disorder across social and demographic covariates. Error bars represent 95% confidence intervals. MOOD = any mood disorder; ANX = any anxiety disorder; SUD = any substance use disorder; STRESS = any stress-related disorder; SCZ = schizophrenia; PERS = any personality disorder

### Patterns of Comorbidity in the All of Us Data

Figure 2 presents the tetrachoric correlations and patterns of comorbidity for each of the six classes of disorders included in the current analysis. In keeping with the All of Us data use requirements, we filtered all cells with fewer than 20 respondents. In terms of correlations, there is significant correlations across all disorders ranging from *r* = 0.43 to *r* = 0.75. In terms of specific patterns of overlap, 57% of the approximately 70,000 diagnoses involved some overlap and 30,798 (~43%) occurred in isolation. Of those with two registered diagnoses (n = 22,797), the vast majority had some configuration of mood and anxiety disorder. Those with three disorders (n = 10,968) mostly had a mood disorder, an anxiety disorder, and either an SUD or stress-related disorder. The last pattern of comorbidity that occurs relatively frequently was among those with a registered diagnosis of a mood, anxiety, substance use, and stress-related disorder. Overall, these patterns of comorbidity demonstrate the fact that psychiatric disorders more frequently occur in conjunction than on their own.

**Figure 2:**
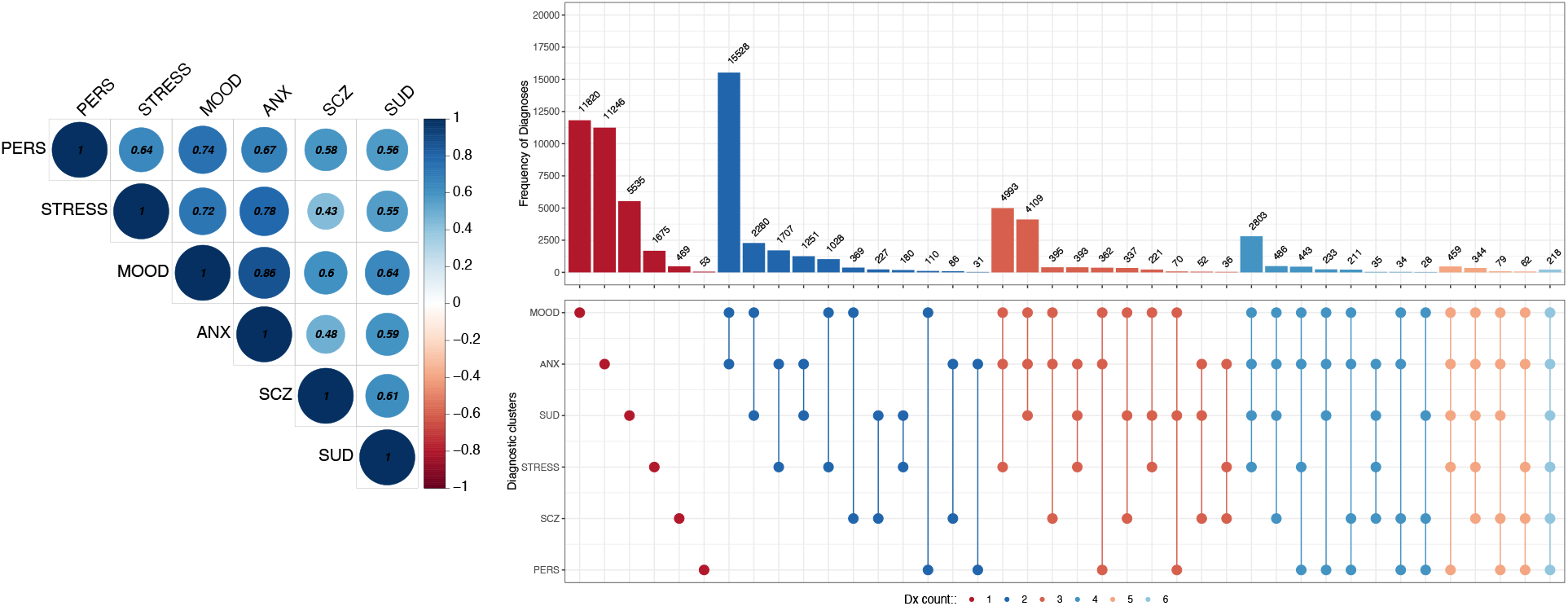
Correlations and Patterns of Comorbidity across Six Classes of Disorders. Tetrachoric correlations (left panel) and overlap between classes of disorders (right panel). All correlations significant at *p* < .001 threshold. Count for each combination of disorder presented at the top of the bar chart in right panel. Bottom right panel denotes the combination of disorders for counts in the above plot. MOOD = any mood disorder; ANX = any anxiety disorder; SUD = any substance use disorder; STRESS = any stress-related disorder; SCZ = schizophrenia; PERS = any personality disorder.

## DISCUSSION

The goal of the All of Us program is to create a resource for health research that can benefit *all* of the population in the United States, with a particular emphasis in improving health equity and representation in medical research. In the current analysis, we provided an overview of the currently available data related to psychiatric and substance use disorders, how these disorders vary across social and demographic factors, and how these disorders cluster together.

In terms of raw prevalence, the disorders captured in the All of Us EHR occur at lower frequency than estimates from nationally representative samples. There are two possible explanations. First, is that survey-based instruments overestimate the prevalence of disorder. However, the more likely explanation is that there is ascertainment bias in All of Us and among individuals seeking treatment for psychiatric disorders more generally. Other biobanks have known issues with ‘healthy participant’ bias^28^. While biobanks provide a useful resource for many medical conditions, household survey approaches may be better able to access hard-to-reach populations, like those with severe mental illnesses. As expected from prior research^7,8,29^, we demonstrated widespread comorbidity between disorders. Overall, these disorders were strongly correlated with one another, though the patterns ranged from very common (mood + anxiety disorders) to less common (4 + disorders).

The pattern of associations across social and demographic factors is similar to those from other randomly drawn samples. Differences in risk across gender identity and sex assigned at birth are generally similar and in line with prior research, whereby women and those assigned female at birth are at increased risk for mood and anxiety disorders, but at lower risk of SUDs^21^. Importantly, All of Us includes a sizeable number of individuals whose identity does not fall into dominant categories in terms of sex assigned at birth, gender identity, and sexual orientation. Persons in these categories were at increased risk virtually each of the classes of psychiatric disorders, similar to prior research^30,31^. The increase prevalence of disorders among these persons is likely the consequence of increased exposure to discrimination and other stressors associated with holding a marginalized identity, similar to those in other marginalized social positions ^32,33^. Prior research has demonstrated that states with more protective policies towards LGBTQ+ persons saw reduced associations between sexual minority status and psychiatric disorder^31^. Further work into policies that can reduce the disparities within these groups is necessary.

In addition to diversity across sex, gender, and sexuality, All of Us also contains substantial diversity across race and ethnicity. Compared to non-Hispanic White participants, those who identify as Black/African American, Hispanic or Latino/a/x, Asian, multiracial, or other racial ethnic category were at *reduced* risk of most disorders, a pattern observed across most psychiatric disorders^34,35^. Given racial health disparities in other medical conditions attributed to racism, racial discrimination, and racial inequality more broadly^36,37^, the opposite direction of association for mental health and psychiatric disorders has proven difficult to explain. It is possible that the items used to assess disorders do not transport well across cultural or racial-ethnic contexts ^38^, and we need disorders/items that accurately capture different racialized experiences^39^. Other possibilities include increased unhealthy behaviors used as a coping mechanism that consequently worsen physical health disparities but mitigate against mental health disparities^40^, or the increased consumption of pharmaceuticals by non-Hispanic Whites that include mental health related side effects^41^, among others. The one exception to the above pattern was in regard to schizophrenia. Like previous findings, the increased prevalence of schizophrenia among those who identify as Black/African American is also a well-documented pattern in social epidemiology^34^. This disparity has been attributed to social antecedents (e.g., racism, lack of access to care)^42^ as well as racial bias in the diagnostic category for schizophrenia itself^43^. Finally, those who are born outside of the US have consistently lower rates of each of the disorders included. These decreased rates could reflect a healthy immigrant effect^44^ or other protective cultural factors among immigrants.

Socioeconomic disparities in all of the disorders are apparent in both education and income. Compared to those with a college degree, all of the other levels of educational attainment are at increased risk of each class of disorder. The gradient in psychiatric disorders is most apparent in household income, with those in the >$25K annually category being at consistently highest risk. These consistent patterns could be due to the increased stress and reduced resources experienced by those at the lower end of the socioeconomic continuum^45–47^, or the reduced upward social mobility of those experience psychiatric disorders (e.g., social drift)^48^, though it is likely a mixture of both.

Our analysis is not without limitations. First, we grouped many different ICD10 codes and diagnoses into larger categories. We did this for simplicity, as the number of available diagnoses in the EHR is tremendous. Future iterations can take a more granular approach for certain diagnostic subcategories. Second, we focused on lifetime diagnoses of disorders. Next steps should also focus on the occurrence of more recent onset disorders and better tease apart time ordering in regard to psychiatric disorders and other health conditions. Future work can also leverage the extensive EHR data to model the course of treatment for specific disorders, and how this may vary across social and demographic contexts.

The All of Us biobank presents a vital resource in better characterizing the etiology of psychiatric conditions and advancing health equity. In the current analysis we have demonstrated the rich data available on psychiatric disorders. Future research will be able to harness the vast genetic, lifestyle, and other social data to better understand the causes and consequences of psychiatric conditions. As we move towards an era of precision medicine, biobanks such as All of Us will be an invaluable resource.

## Data Availability

All of Us data are accessible through the All of Us Researcher Workbench. For more information on how to access visit: https://workbench.researchallofus.org/

## ACKNOWLEDGMENTS

The *All of Us* Research Program is supported by the National Institutes of Health, Office of the Director: Regional Medical Centers: 1 OT2 OD026549; 1 OT2 OD026554; 1 OT2 OD026557; 1 OT2 OD026556; 1 OT2 OD026550; 1 OT2 OD 026552; 1 OT2 OD026553; 1 OT2 OD026548; 1 OT2 OD026551; 1 OT2 OD026555; IAA #: AOD 16037; Federally Qualified Health Centers: HHSN 263201600085U; Data and Research Center: 5 U2C OD023196; Biobank: 1 U24 OD023121; The Participant Center: U24 OD023176; Participant Technology Systems Center: 1 U24 OD023163; Communications and Engagement: 3 OT2 OD023205; 3 OT2 OD023206; and Community Partners: 1 OT2 OD025277; 3 OT2 OD025315; 1 OT2 OD025337; 1 OT2 OD025276. In addition, the All of Us Research Program would not be possible without the partnership of its participants.

